# Mixed Methods Family Centered Study of Pain Experience in Non-Hispanic White and Black Children

**DOI:** 10.1101/2024.03.20.24304546

**Authors:** Julia Kumar, Dylan Atkinson, Adaora Chima, Laura McLaughlin, Rajvi Parikh, Peter Mende-Siedlecki, Monica Mitchell, Vidya Chidambaran

## Abstract

**Background and Objectives:** Although racial disparities in pediatric pain have been described, family-centered research is lacking. This mixed-methods study aimed to understand race-specific factors impacting acute pain experiences in Non-Hispanic White (NHW) and Black (NHB) children.

**Methods:** NHW and NHB children (aged 8-17) (n=19 each) with a recent acute pain experience, and their caregivers were recruited. The following domains were assessed in children (deprivation, ethnic identity, pain, psychosocial, pain coping, resilience) and caregivers (education, income, Racial and Ethnic Microaggressions Scale (REMS), Adverse Childhood Events (ACEs), Adult Response to Child’s Symptoms). Questionnaire measures were compared between groups using simple statistical tests. Fifteen dyads participated in focus groups. Thematic codes underlying pain experiences were identified.

**Results:** NHB children had similar pain/psychosocial characteristics but differed in ethnic identity (2.7 (0.5) vs. 2.2 (0.5); P=.002), deprivation index (0.4 (IQR 0.3-0.5) vs. 0.3 (IQR 0.2, 0.3), P=.007) and pain coping efficacy (8.6 (0.4) vs. 9.8 (0.5), P=.045) from NHW children. NHB caregivers scored higher on REMS sub-scales, ACEs (0 (0,1) vs 0 (0,0); P=.02) and Protection (1.9 (0.6) vs. 1.6 (0.5); P=.03) behaviors in response to child pain. NHB and NHW participants endorsed satisfaction with pain experiences, opioid avoidance, and stoicism. Unlike NHW participants, NHB reported barriers related to trust, discrimination, and access.

**Conclusions:** Racial differences in acute pain experiences suggest healthcare providers need to reinforce trust and consider underlying cultural and pain coping differences when treating pain in NHB children. Findings emphasize that family-centered and systems-based approaches are important for equity in pediatric pain.

**Article Summary:** This mixed-methods family-centered study identifies common themes, systems-related barriers and pain coping efficacy differences underlying pain experiences of Black and White children.

*What’s Known on This Subject:* Inequities exist in post-surgical pain and pain management of Black and White children.

*What This Study Adds:* Family perspectives regarding pain experience of NHB and NHW children highlight inter-group differences in stressors, barriers, pain coping and parent responses to child’s pain. Findings underscore the importance of family engagement and addressing systemic barriers to improve pain equity.

**Contributors Statement:** Chidambaran conceptualized and designed the study, conducted focus groups, coordinated and supervised data collection, analyses, and critically reviewed and revised the manuscript for important intellectual content.

Mitchell conceptualized and designed the study, critically reviewed and revised the manuscript for important intellectual content.

Kumar and Atkinson conducted subject recruitment, collected data, drafted the initial manuscript, critically reviewed and revised the manuscript, and designed the data collection instruments in REDCap, and analyzed the data.

McLaughlin, Parikh and Chima participated in conduct of focus groups and maintained regulatory aspects of the study, critically reviewed and revised the manuscript

Mende-Siedlecki helped design the study, critically reviewed and revised the manuscript. All authors approved the final manuscript as submitted and agree to be accountable for all aspects of the work.

**Data Sharing Statement:** Deidentified individual participant data (including data dictionaries) will be made available, in addition to study protocols, the statistical analysis plan, and the informed consent form. The data will be made available upon publication to researchers who provide a methodologically sound proposal for use in achieving the goals of the approved proposal. Proposals should be submitted to.

## Introduction

Despite advances in our understanding of pain and pain management, pain continues to be prevalent, underrecognized, and frequently undertreated in pediatric populations.^1^ Importantly, pain does not impact everyone equally. Differences in risk and susceptibility to pain have been attributed to race and ethnicity. This has been shown to occur in various settings, both pediatric and adult, whether it is related to chronic pain-related conditions,^2^ pain management in the emergency department,^3^ or opioid requirements after surgery.^4, 5^ Further, stereotyping and dogma about risky health behaviors also impact racial disparities in treatment.

In 2003, the Institute of Medicine (IOM) report “Unequal Treatment,” laid out a broad theoretical framework to address the root causes of health care disparities within three categories: the patient, the health process, and the health system.^6^ The American Academy of Pediatrics, the National Institutes of Health and others have prioritized Diversity, Inclusion and Equity in Medicine and Research in recent years.^7, 8^ However, research in areas such as pediatric pain are scarce despite evidence that postsurgical pain and disabilities disproportionately affect children from marginalized racial and ethnic populations.^2^ Since childhood and adolescence are sensitive periods during which daily life challenges continuously shape brain circuitry and adaptive health behaviors, environmental exposure to higher stress, including adverse childhood experiences and low socioeconomic status are expected to impact pain experiences several years after the onset of events.^9^ Cultural identity and beliefs, parental attitudes, and pain coping styles ^10, 11^ may also influence the pain experiences of children and adolescents. Therefore, family-centric approaches are critically needed^12^ to enable targeted strategies to reduce bias and inform future actions.^13^ The purpose of this novel mixed-methods study was to gain a more empathic understanding of acute pain experiences in Non-Hispanic Black (NHB) and White (NHW) cohorts, by engaging the family. Goals were to describe common and race-specific characteristics and themes in pain-related domains (satisfaction, pain assessment and management, stressors, barriers, caregiver responses, pain coping and resilience).

## Methods

This nested, mixed methods study was conducted in a tertiary pediatric institution, as part of a larger study recruiting NHB, NHW, Asian and Hispanic families. The study was approved by Cincinnati Children’s Hospital institutional review board. Written informed consent was obtained from caregivers (parents or legal guardians for minor subjects) and written assent from children aged 11 and above.

### Participant Recruitment

English-speaking children (8-17 years of age) who self-identified as Non-Hispanic Black or White with a recent acute pain experience were recruited. Acute pain experience was defined and satisfied as follows; pain from injury, surgery, or other etiology that lasted <14 days, for which they required pain management at a medical facility within the previous two years. Primary caregivers who directly cared for the child during this experience were also recruited. Potential participants were identified from electronic medical records or via dissemination of information through flyers and at community events. Those that responded to flyers/community outreach that did not meet recruitment criteria were excluded. Children with developmental delays, chronic/cancer pain and currently hospitalized, were excluded. Participants (children and caregivers) were not included if they indicated inability or unwillingness to provide feedback on the recent pain experience. Prior to consenting participants, study team members discussed details of the study over the phone and provided the option of only answering questionnaires if unable to participate in focus groups. The dyad was compensated with $40 if they only answered questionnaires and $100 if they participated in focus groups.

### Questionnaire administration

Following informed consent/assent, participants were sent questionnaires to the caregivers’ and/or child’s email (as preferred by participants) through REDCap database. Both caregivers and children were responsible for independently completing their assigned questionnaires. Caregivers were instructed to notify the study team if their child had difficulty completing their questionnaires. Questionnaires did not need to be completed prior to participating in a focus group; however, completion of caregiver and child questionnaires was required before participants could be compensated. Demographic information, details of the pain episode (reason, nature of pain, treatment facility), and residential address were obtained.^14^

All the questionnaires and the corresponding domains assessed are described in Table 1. Child domains assessed were multigroup ethnic identity measure (MEIM),^14^ pain,^15^ functional disability,^16^ quality of life,^17^ psychosocial measures^18^ (anxiety, depression, family relationships, psychological stress), pain coping,^19^ exposure to traumatic events and Adverse Childhood Experiences (ACEs), ^20^ mindfulness,^21^ and resilience.^22^ Caregivers were asked about their highest education, annual income, pain catastrophizing,^23^ ACEs.^20^ perceived stress,^24^ Racial and Ethnic Microaggression,^25^ and Adult response to Children’s Symptoms^26, 27^ which described parental behaviors in response to their child’s pain.

**Table 1:**
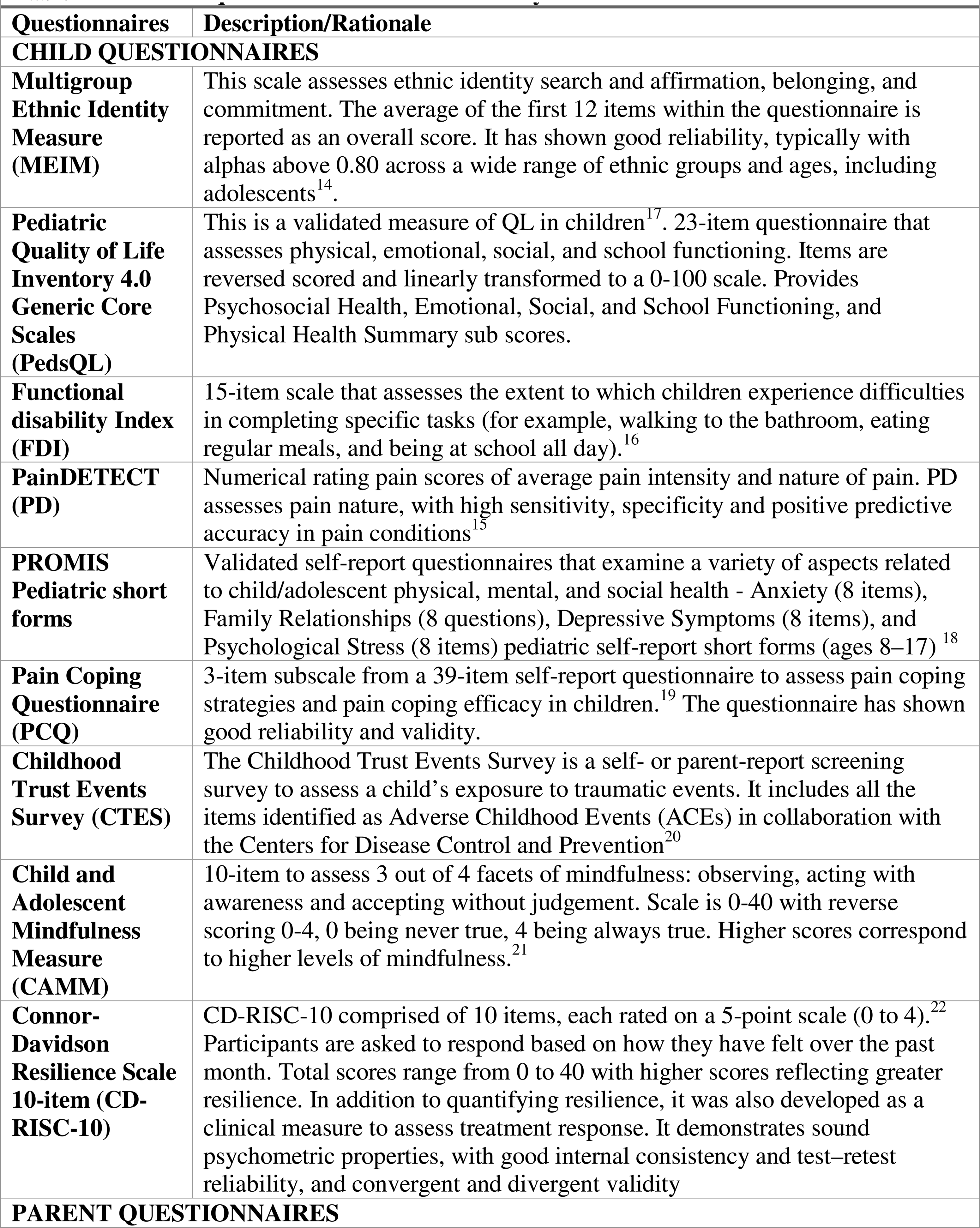
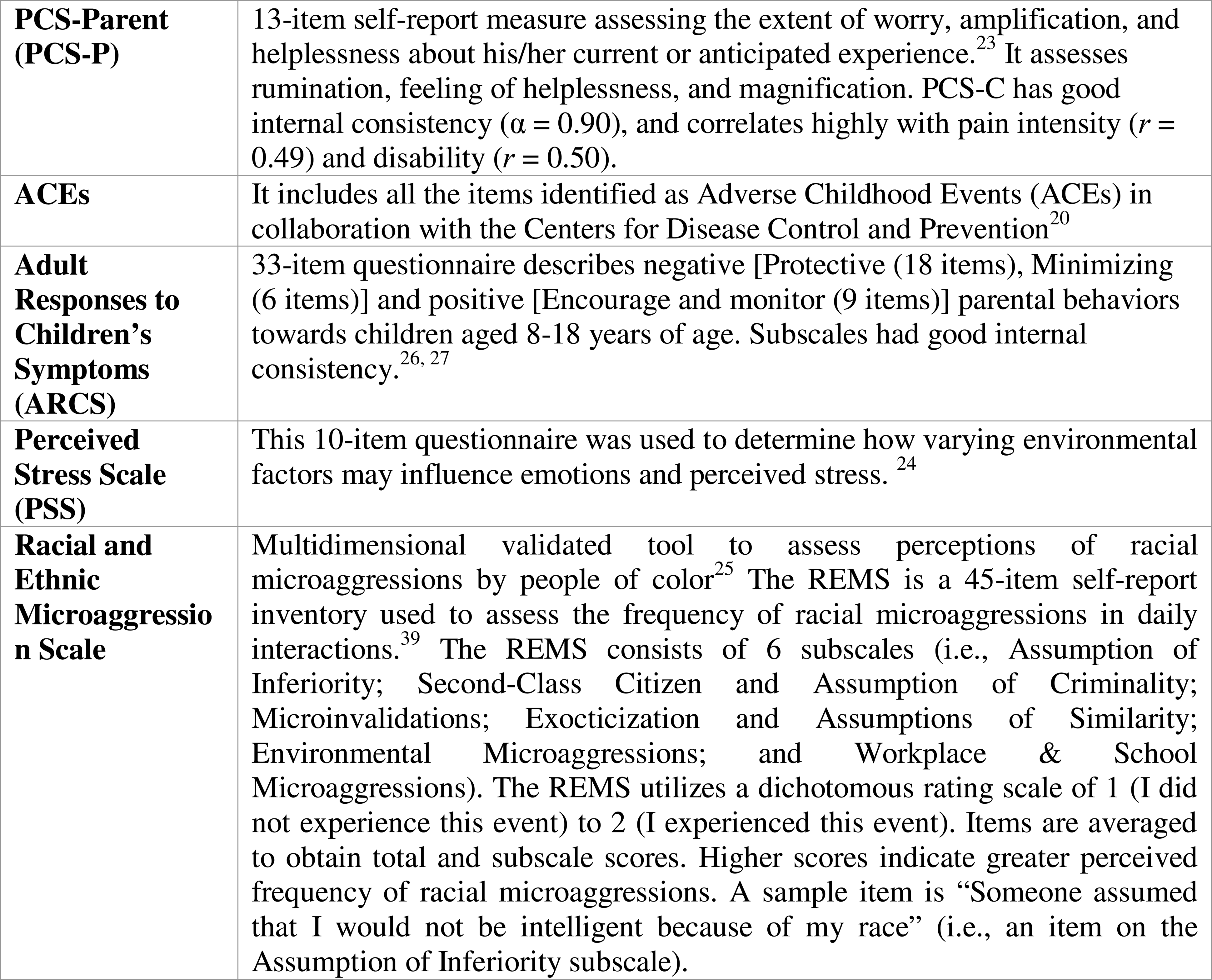
Validated questionnaires used in the study.

### Focus Groups

Focus groups were conducted in accordance with COREQ criteria for qualitative research.^21^ Each group consisted of 4-6 race-homogenous, child-caregiver dyads. Virtual sessions were hosted via Zoom meeting software, lasting up to 90 minutes. They were led by two trained members of the project team using a standardized interview script to assess several domains of interest (Supplementary Figure 1). After introductions, statement of rules for the FG, discussions first engaged children (about 40 minutes) followed by caregivers (40 minutes). This sequence was followed so the children could be relieved earlier. Caregivers were advised not to answer for their children but were allowed to prompt or explain the question if need be.

### Data Analysis

We calculated summary statistics of variables and questionnaire scores and compared NHB and NHW cohorts, using simple statistical tests. Focus groups were recorded and transcribed orthographically, reproducing all spoken words and sounds, using NVivo software (version 1.6.1, QSR International Pty Ltd., NVivo, released March 2020). After each focus group, facilitators documented and discussed initial general impressions. Transcripts were de-identified and edited for brevity, removing words/clauses not essential for understanding the overall meaning of the data. Subsequently, transcribed focus group data were reviewed and independently coded. Codes were then compared, reconciled, and used to extract sub-themes based on induction and prior knowledge utilizing the 6 steps described by Braun and Clarke^28^ (i.e. getting familiar with the data, creating initial codes, identifying themes, reviewing and refining themes, defining and naming themes, and producing the report). We used triangulation methods for mixed-methods integration to leverage explanatory sequential design of quantitative data collection.^29^ The original goal was to recruit 25-30 dyads, but recruitment was considered sufficient when a new FG did not further enrich the themes. Prior research has suggested that two thirds of themes can be identified from the first FG and that more than 80% of themes can be generated after 2-3 FGs.^30^

## Results

19 NHB and NHW dyads were recruited, of which two NHB dyads were recruited through community flyers. Four dyads opted to only fill questionnaires and 15 participated in four focus groups in each cohort. All participants completed questionnaires except one NHW child. Several attempts to secure completed questionnaires from this participant were unsuccessful. The data from the corresponding caregiver’s questionnaires is still included in data analysis. Demographics and questionnaire measures are summarized in Table 2. The cohorts had similar age and sex distribution. NHB and NHW cohorts had similar pain etiology, location of pain management (hospital admission in 61% of NHB and 71% of NHW), pain and psychosocial measures. NHB children identified more strongly with their ethnicity than NHW children (P=.002). NHB had lower pain coping efficacy (P=.045), although pain coping behavior scales were similar.

**Table 2:**
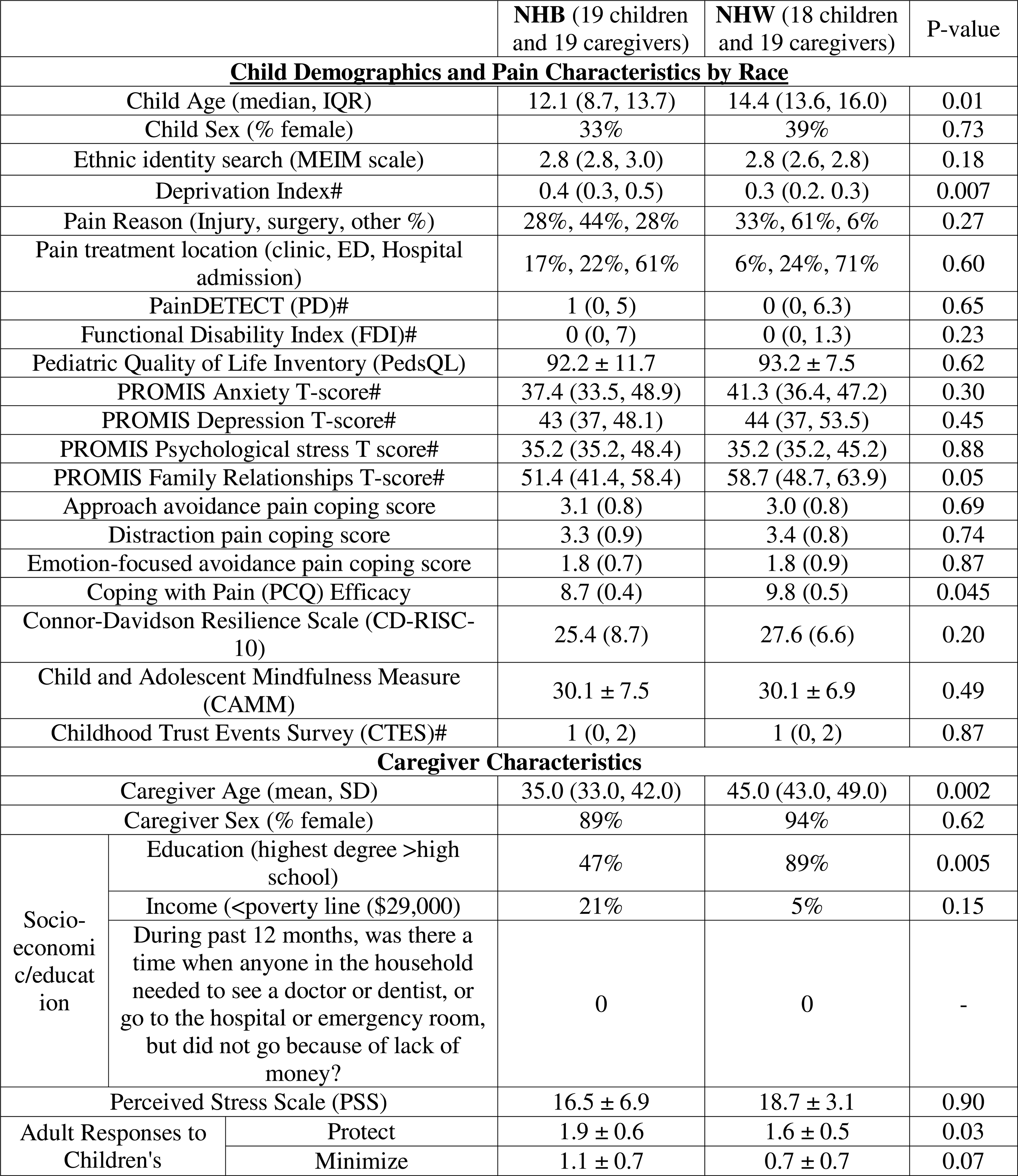
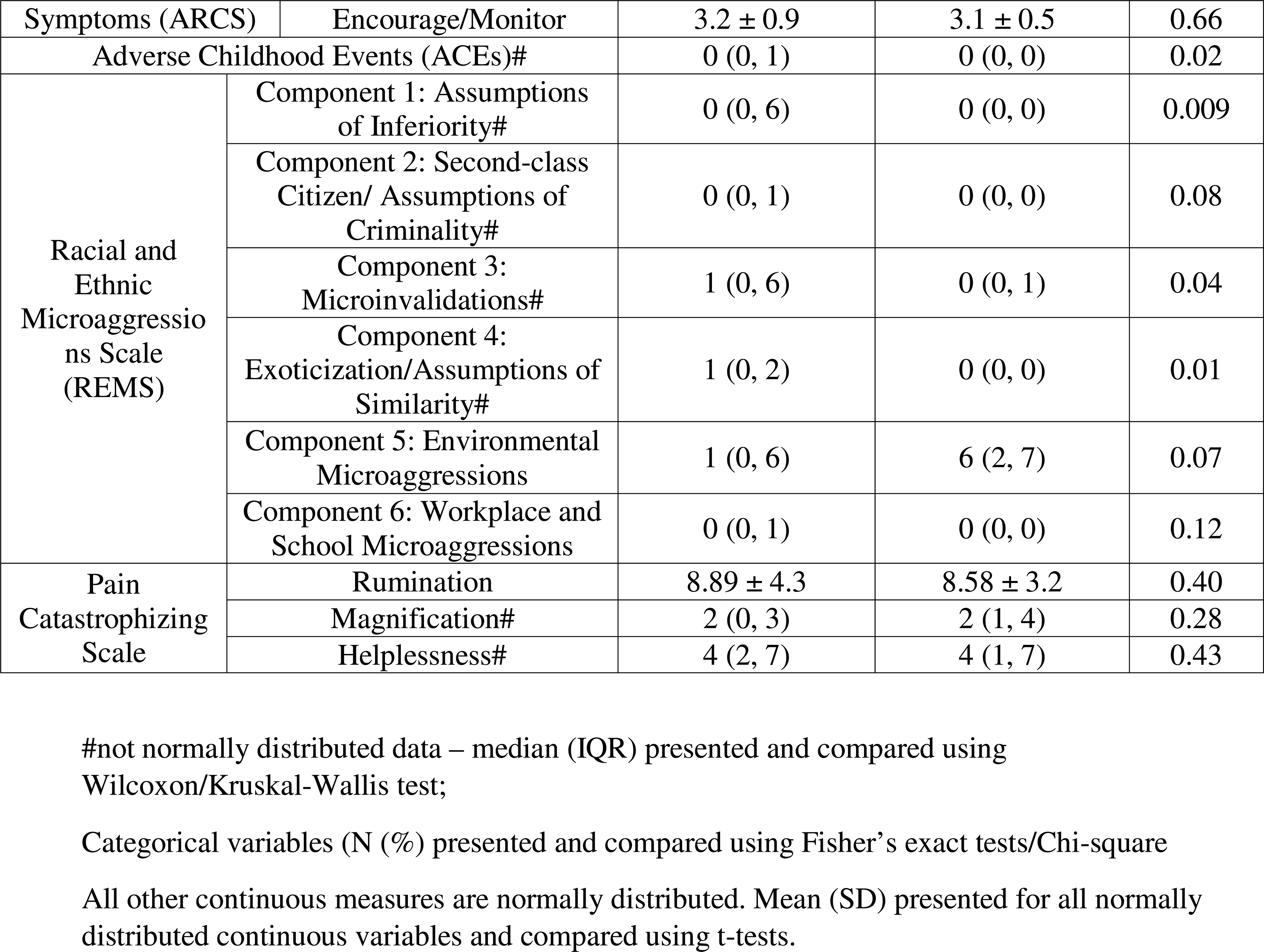
Comparison of child-caregiver characteristics in acute pain settings in Non-Hispanic black (NHB) and White (NHW) children.

NHW caregivers had a higher proportion of college graduates (P=.005) and annual income>$29,000 (P=.150) compared to NHB caregivers. NHB caregivers had significantly higher scores for several subscales on the Racial and Ethnic Microaggressions Scale, namely, Assumptions of Inferiority, Microinvalidations and Similarity; and higher ACE scores (P=.02). NHB caregivers used Protection (P=.03) and Minimization (P=.07) behaviors to help children cope with pain more than NHW caregivers.

Below we present key themes based on thematic analyses of focus groups (FG). Supporting quotes are provided in Table 3. Quotes are referenced as BC or WC for NHB and NHW caregiver quotes, and Bc and Wc for NHB and NHW children quotes respectively. The frequency with which NHB and NHW cohorts endorsed positive and negative sentiments coded to trust, satisfaction, access and barriers, are presented in Figure 1.

**Figure 1.**
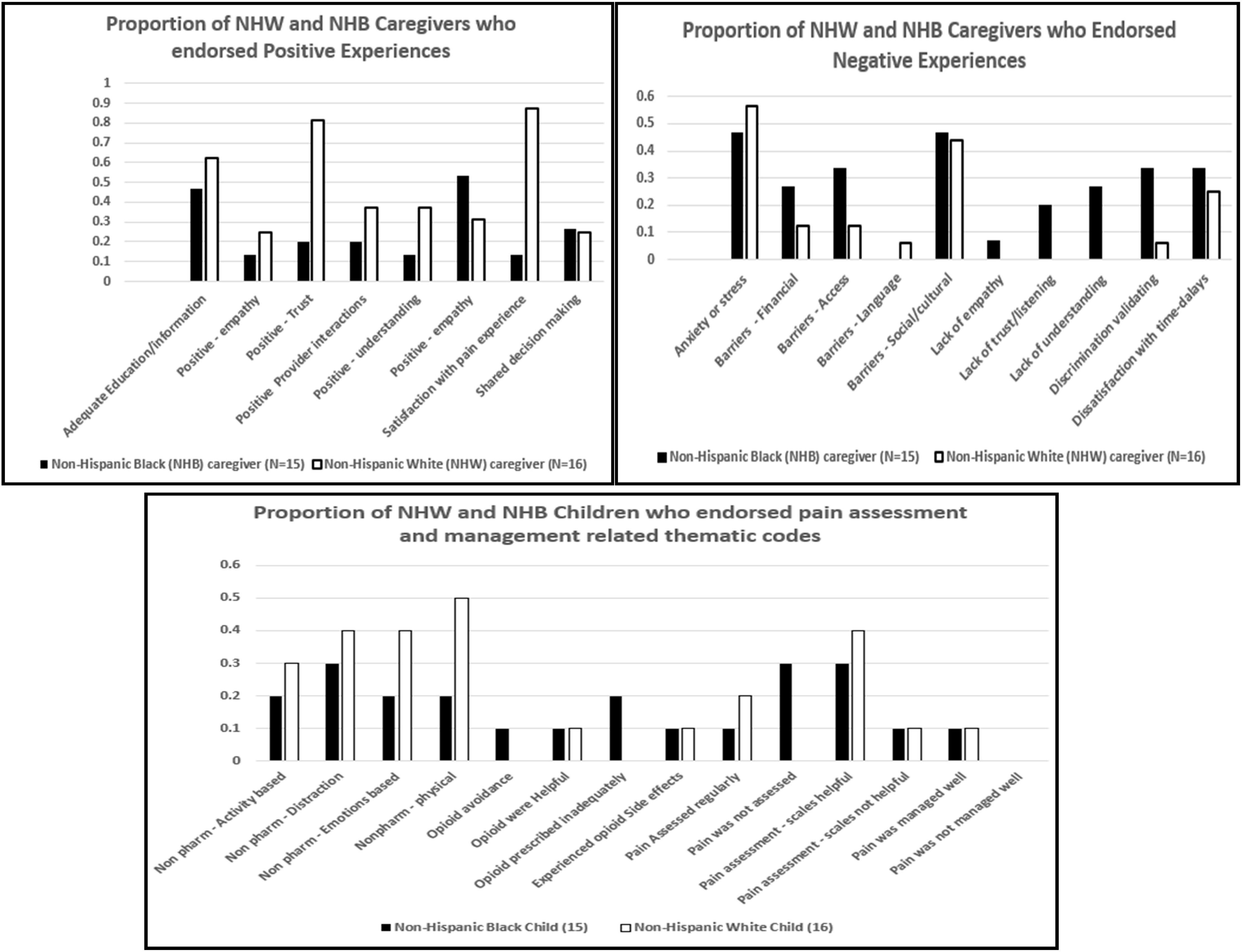
Representation of proportion of NHB and NHW caregivers and children contributing to thematic codes. The y-axis represents the fraction of participants endorsing the sentiment. The black filled bars in both upper and lower panel represent Black cohort and the white unfilled bars the White cohort. The upper panel depicts the positive (left side) and negative sentiments (right) sentiments endorsed by caregivers and the lower panel depicts the proportion of NHB and NHW children who endorsed codes related to use of non-pharmacological methods and opioids for pain management, pain assessment and management during focus groups. Non pharm: non-pharmacological methods of pain management.

**Table 3:**
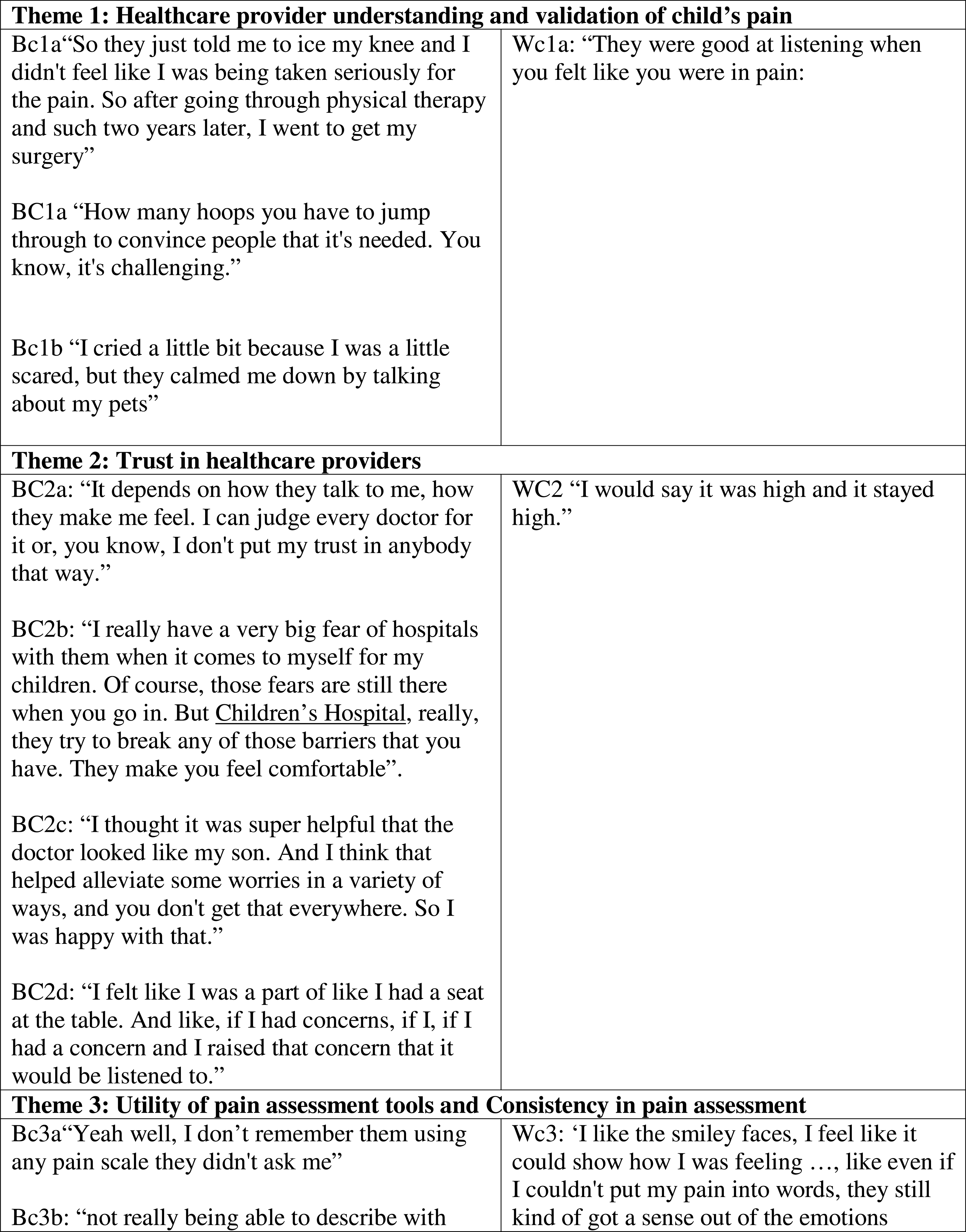
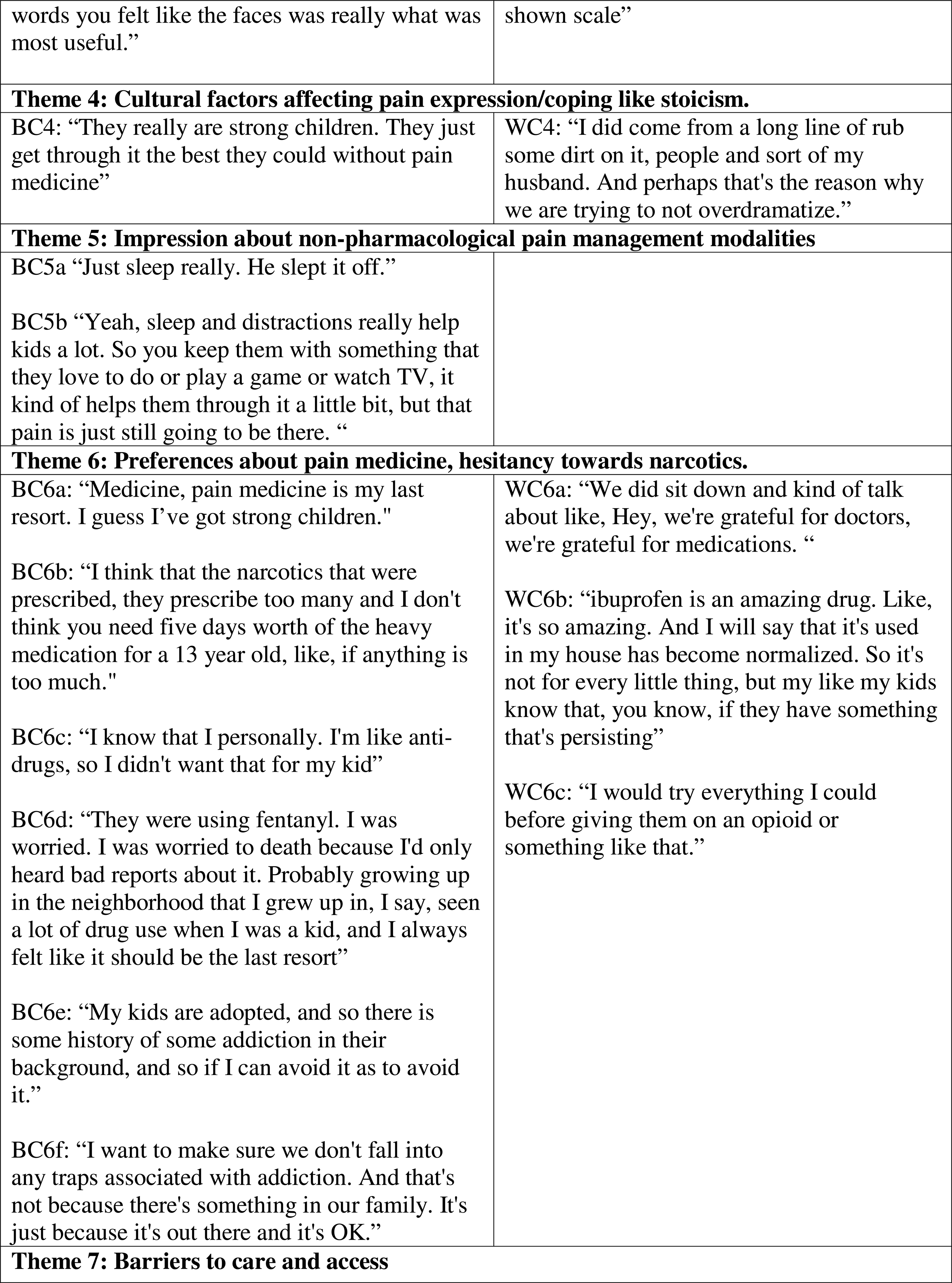
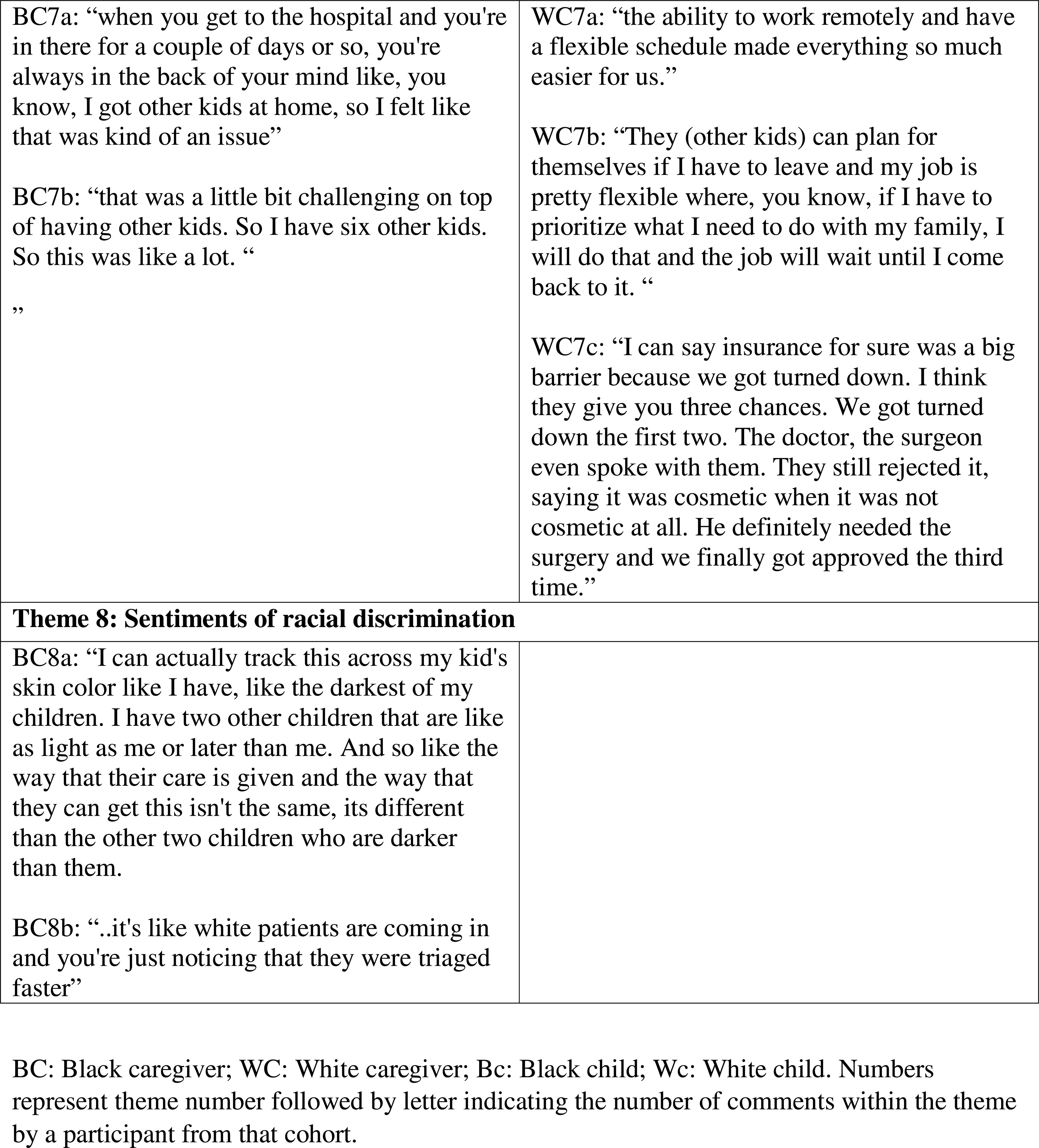
White and Black children and caregiver quotes lending credence to the themes based on coding and analyses.

### Theme 1: Healthcare provider understanding and validation of child’s pain

Both NHB and NHW cohorts expressed satisfaction with how their child’s pain was managed. However, about 27% of NHB caregivers (4/15) expressed negative sentiments consistent with the thematic code “pain not managed well,’ compared to one NHW caregiver (Figure 1).

Additionally, about 27% of NHB caregivers (4/15) expressed *lack of validation* of their pain from the healthcare providers, but none in the NHW (Figure 1; BC1a, Bc1a). That said, children in both cohorts endorsed receiving empathetic care (Table 3; Wc1a, Bc1b).

### Theme 2: Trust in healthcare providers

NHW caregivers expressed positive sentiments of trust in their providers (WC2). Three NHB caregivers explicitly expressed lack of trust or hesitancy about trusting their providers (BC2a). Some NHB caregivers felt more trust in providers following a positive experience in their child’s pain management (BC2b). Importantly, one NHB caregiver reported a higher level of trust and relief in being better understood when the healthcare provider was also Black (BC2c) Another positive comment from a NHB caregiver suggested being part of shared decision-making increased trust. (BC2d)

### Theme 3: Utility of pain assessment tools and Consistency in pain assessment

The majority of NHW children remembered being asked to report their pain on a scale, four NHB children reported never having their pain assessed using a scale. Overall, both groups considered the numeric pain scale to be useful and effective, except a couple of instances in Black children (Bc3a, Bc3b). Both NHB and NHW children (Wc3) and caregivers felt that the combination of numbers and faces/images was effective for children. Children from both cohorts felt being prompted by providers with adjectives, such as “achy,” “dull,” or “stabbing,” was useful when asked to describe their pain. Figure 1 (lower panel) depicts the children’s sentiments.

### Theme 4: Sociocultural factors affect pain expression/coping

Themes of stoicism were identified in both cohorts. Both NHB and NHW children cited “toughing out the pain,” which was further endorsed by NHB and NHW caregivers (BC4a and WC4a). Both caregiver cohorts also mentioned social support and faith helped them cope with the difficult experience.

### Theme 5: Impression about non-pharmacological pain management modalities

NHB and NHW children described the benefits of being able to distract themselves from pain. They mostly cited playing video games and spending time with family or friends. There were subtle differences between cohorts in the use of physical methods such as ice, elevation, and massage. Interestingly, children in both groups described “sleeping off their pain.” (BC5a and b).

### Theme 6: Opinions about analgesics and hesitancy towards narcotics

NHW caregivers were more accepting of non-opioid medication use, while NHB caregivers preferred to avoid even over-the-counter medications (WC6a-b and BC6a-c). However, when asked about opioid analgesia, caregivers from both groups demonstrated hesitancy (9/15 NHB and 8/15 NHW) with a strong preference for reserving opioids for last resort scenarios as they associated opioids with addiction (WC6c and BC6b-f). While some expressed these concerns based on news articles they had read, several noted experiences with family members and neighbors.

### Theme 7: Barriers to care and access

Several NHB caregivers noted difficulty balancing their child’s care with existing responsibilities, namely inflexible work schedules or caring for additional children (BC7a, BC7b). Conversely, several NHW caregivers expressed greater ease in handling their child’s care due to flexible work schedules (WC7a, WC7b). Two of 15 NHB caregivers identified transportation as a challenge during their child’s pain care management. Furthermore, several caregivers, both NHB and NHW, commented that negotiating with health insurance was particularly frustrating (WC7c).

### Theme 8: Sentiments of racial discrimination

In both NHB and NHW cohorts, the question of whether racial discrimination influenced pain experience was typically answered quickly with “no” or met with silence. Further prompting elicited considerable feedback from NHB caregivers, particularly to the question “Do you feel that people of different races and incomes are treated differently in these situations?” They shared sentiments that reflected anxiety over how the racial identity of their child was perceived in healthcare settings (BC8a). One NHB caregiver stated unfairness in access to medical care in the ED leading to a longer wait compared to White patients (BC8b), and another mother reported her child was given less than adequate pain medication.

## Discussion

This study utilized qualitative and quantitative research methodology for a family centric understanding of acute pain experiences among children from NHW and NHB racial backgrounds. The data from questionnaires corroborated themes that emerged from focus groups, confirming the differences in life experiences between NHW and NHB caregivers, the unique barriers faced by NHB cohort and some mutual themes challenging to both cohorts, in dealing with acute pediatric pain experiences.

The diathesis-stress hypothesis states that cumulative lifetime stress can contribute to worse pain experiences.^31^ Black Americans have been shown to display more “weathering” and higher levels of allostatic load compared to White populations, with worse pain-related outcomes in adolescents.^32^ In our study, NHB caregivers reported higher ACEs, racial discrimination, lower education level, and higher socioeconomic constraints. They reported additional responsibilities and work restrictions that made it difficult to care for their child. Assumptions of inferiority, microinvalidation and exoticization scores were higher in the NHB cohort compared to NHW. These refer to perception that one is considered as ‘less’ or expressions of surprise at personal achievements and bias in validating pain expressions. In fact, NHB children expressed lack of validation of their pain and need to overemphasize their pain in order to receive treatment. These and other experiences of discrimination further erode trust in the doctor/patient relationship. They are aligned with previous reports that providers held implicit bias that Black Americans were more pain-tolerant, contributing to lack of cultural sensitivity and compassion^33^. Thus, providers may evaluate NHB patients more negatively than they do similar NHW patients, as they perceive them as more likely to participate in risky health behaviors and may be less willing to prescribe them pain medications and narcotic medications.

In caring for pediatric patients, the clinical team often depends on caregivers to ascertain the presence and character of pain, and the impact of interventions.^34^ These advocates may subconsciously confer their personal perspective on the patient’s experience. It is possible that their lifetime experiences of the caregivers inadvertently lead NHB caregivers to develop more protective responses while caring for their child in pain.^35^ This is shown by our findings of higher protective behaviors in NHB caregivers. In addition, we found lower pain coping efficacy in the NHB children. A higher tendency to engage in protective behaviors when reacting to their child’s pain (as in NHB caregivers) has previously been linked to maladaptive child outcomes, including higher disability and somatic symptoms.^36^ It is possible that different racial, ethnic and/or cultural backgrounds have differing coping responses. These are novel findings with potential implications for pediatric pain management strategies in racially diverse groups. Thus, tailored interventions may be indicated for improved pain coping efficacy and one size may not fit all.^37^

Our study did not find psychosocial differences between NHB and NHW caregivers or children, unlike studies that suggest higher degrees of catastrophizing in African American patients.^38^ We found stoicism and opioid aversion expressed by both cohorts. NHB caregivers echoed pride in their children being resilient and strong-willed enough to endure their pain with little complaint. NHB children also mentioned choosing to “sleep off pain.” Children in both cohorts found spending time with family, friends, and utilizing other forms of distraction effective in dealing with pain. Our study also corroborated findings from previous studies that mention prayer as a significant race-specific method of pain coping.^38^ Hesitation to use opioids in managing pediatric pain was demonstrated in over 50% of NHW and NHB participants. The common avoidance of opioid use fueled by opioid addiction fears suggest more education is needed so these fears do not prevent optimal pediatric pain management.

This study is unique in its patient-centered approach, with standardized FGs conducted by both a trained moderator and a pain expert. Nevertheless, differences between study groups may limit the conclusions. First, although in broad terms, pain experiences were similar between groups (post-trauma or post-surgical pain), differences in injury mechanism and operative procedure may have influenced perceived pain. For example, a higher proportion of the NHW cohort experienced major surgery related pain (such as pectus repair), compared to the NHB cohort. While this would be expected to result in more severe pain in this cohort, they perceived the experience as less stressful. This could be because these surgeries were conducted in a tertiary pediatric center, and pain managed by a dedicated team using standardized multimodal pain management approaches. Secondly, a potential limitation is the possible influence of the moderators’ race and sex on the participants’ feedback. FGs were run by race diverse and congruent moderator pairs whenever possible. Perceived racial congruence with FG moderators may have increased or decreased participants’ comfort with sharing especially negative opinions. Thirdly, although we obtained consistent feedback from children of all ages, children sometimes misinterpreted the question (for example, misinterpreting a question about pain medication as their experience with general anesthesia administration). Fourthly, while presence of caregivers during pediatric FG was deemed necessary for the comfort of the participants, they sometimes would prompt the responses despite instructions not to do so. Finally, families of Hispanic ethnicity and other races were not included as part of this nested study to enable focus on NHB specific factors influencing pain experiences.

## Conclusions

Equity in healthcare has long been a challenging goal to attain in different sectors of healthcare, including pain management. Our mixed methods study confirms racial differences exist in experiences of acute pediatric pain and emphasizes the need for family centered and systems-based approaches to improve equity in pediatric pain care. Increasing representation of diverse racial groups within the healthcare workforce would be an important step in increasing trust in the system and introducing tailored strategies to enhance pediatric pain coping. In addition, racial differences in underlying stress exposures should also be considered by healthcare providers and policymakers while co-creating strategies to address racial bias in pain management.

## Data Availability

All data produced in the present study are available upon reasonable request to the authors.

## Acknowledgements

We would like to acknowledge Carlos Ortiz, Summer Undergraduate Research Fellow in Chidambaran Lab and Jennifer Allen, Division of Behavioral medicine and clinical psychology, Department of Pediatrics, Cincinnati Children’s Hospital, Cincinnati, OH for their help with setting up data acquisition instruments through REDCap and subject recruitment (Ortiz).

## Abbreviations

NHW: Non-Hispanic White
NHB: Non-Hispanic Black

